# If you build it, will they use it? Use of a Digital Assistant for Self-Reporting of COVID-19 Rapid Antigen Test Results during Large Nationwide Community Testing Initiative

**DOI:** 10.1101/2022.03.31.22273242

**Authors:** Carly Herbert, Qiming Shi, Vik Kheterpal, Chris Nowak, Thejas Suvarna, Basyl Durnam, Summer Schrader, Stephanie Behar, Syed Naeem, Seanan Tarrant, Ben Kalibala, Aditi Singh, Ben Gerber, Bruce Barton, Honghuang Lin, Michael Cohen-Wolkowiez, Giselle Corbie-Smith, Warren Kibbe, Juan Marquez, Jonggyu Baek, Nathaniel Hafer, Laura Gibson, Laurel O’Connor, John Broach, William Heetderks, David McManus, Apurv Soni

## Abstract

**Importance:** Wide-spread distribution of rapid-antigen tests is integral to the United States’ strategy to address COVID-19; however, it is estimated that few rapid-antigen test results are reported to local departments of health.

**Objective:** To characterize how often individuals in six communities throughout the United States used a digital assistant to log rapid-antigen test results and report them to their local Department of Health.

**Design:** This prospective cohort study is based on anonymously collected data from the beneficiaries of The Say Yes! Covid Test program, which distributed 3,000,000 rapid antigen tests at no cost to residents of six communities between April and October 2021. We provide a descriptive evaluation of beneficiaries’ use of digital assistant for logging and reporting their rapid antigen test results.

**Main Outcome and Measures:** Number and proportion of tests logged and reported to the Department of Health through the digital assistant

**Results:** A total of 178,785 test kits were ordered by the digital assistant, and 14,398 households used the digital assistant to log 41,465 test results. Overall, a small proportion of beneficiaries used the digital assistant (8%), but over 75% of those who used it reported their rapid antigen test results to their state public health department. The reporting behavior varied between communities and was significantly different for communities that were incentivized for reporting test results (p < 0.001). In all communities, positive tests were less reported than negative tests (60.4% vs 75.5%; p<0.001).

**Conclusions and Relevance:** These results indicate that app-based reporting with incentives may be an effective way to increase reporting of rapid tests for COVID-19; however, increasing the adoption of the digital assistant is a critical first step.

## Introduction

Rapid antigen home-tests for COVID-19 are an important part of the Federal Government’s strategy for COVID-19 to expand testing access and availability throughout the United States.^1^ However, the distribution and scale-up of rapid home-tests for COVID-19 has been inconsistently accompanied by standard public health reporting mechanisms, challenging the ability to monitor rates of COVID-19 testing. It is important to understand more about individuals’ reporting choices, to create an optimal system for self-testing and surveillance. This study characterized how often individuals in six communities logged their home-test results through a digital platform and patterns of result reporting to state Departments of Health (DoH).

## Methods

### Say Yes! Covid Test (SYCT!) Intervention Communities and Procedures

The Say Yes! Covid Test (SYCT!) program, a partnership between the National Institutes of Health (NIH) and the Centers for Disease Control and Prevention (CDC), distributed over 3,000,000 self-tests to six communities across the United States from April to October 2021.^2,3^ More details about the SYCT! intervention can be found elsewhere.^4,5^ This analysis included data from six communities that finished test distribution before December 2021 and allowed users to report rapid antigen test results to the state department of health through a digital assistant: Louisville, KY; Indianapolis, IN; Fulton County, GA; O’ahu, HI; Ann Arbor/Ypsilanti, MI; and Chattanooga, TN. Test kits were distributed by online ordering and direct shipment to residents’ homes (direct-to-consumer or “DTC”) or local pick-up at community sites. Each household was restricted to ordering one test kit. Test kits in Kentucky, Indiana, Georgia, and Hawaii contained eight rapid home-tests, while those in Michigan and Tennessee contained 25 tests. A $25 gift card incentive was offered to participants in Indiana and Kentucky if they reported at least one test result to their state DoH through the digital assistant. The incentive was also offered in Georgia and Hawaii starting on October 4^th^. This study received non-research determination by the University of Massachusetts Chan Medical School Institutional Review Board.

### Data collection

An optional online platform and accompanying app, developed by CareEvolution LLC, was launched with the SYCT! intervention as a platform for DTC orders, logging test results, and reporting results to the state DoH. All features of the digital tool were freely available and stored without personal identifiable information. The log feature allowed individuals to document their test dates and results for their records. Individuals were given the option to report each logged test to their state DoH through the digital assistant. For logged tests, test date, result (positive, negative, or invalid), and reporting decision (report or no report) were included in a data feed accessible to CareEvolution. For this analysis, reported tests included those reported with personal identifiable information or anonymously. Tests logged in the digital assistant from April 1, 2021 to January 12, 2022 were included in the analyses. Residents of Tennessee were unable to report tests to the DoH until June 24, 2021, so data before this point was excluded from reporting analyses.

### Analyses

Total DTC orders and digital assistant users were calculated by community. The percentage of logged tests reported to the DoH by community was displayed graphically using R 4.1.1.^6^

## Results

### Distinct users for Logging Test Results

In total, 178,785 households ordered test kits through the digital assistant, and 14,398 households used the digital assistant to log 41,465 test results (Table 1). In Hawaii and Georgia, 100% and 66.7% of test kits were distributed before the onset of incentivization, respectively. Of the six intervention communities, Michigan had the greatest proportion of digital assistant users (23.5%) out of total online orders. Overall, 8.1% of individuals who used the digital assistant to order tests opted to log one or more test results on the platform. While the median number of tests logged by participants was one test, a small number of participants (3.1%) logged upwards of 15 test results in the digital assistant.

**Table 1:** Users of MyDataHelps and Tests Reported by Intervention Community.

| Community | Intervention Start Date: | Total Kits Distributed | Digital Direct-to-Consumer Test Kit Orders<sup>a</sup> | Users of the Digital Assistant Test Log | Total Tests Logged in Digital Assistant | Median Test Results Logged per User | Total Tests Reported to Department of Health |
| --- | --- | --- | --- | --- | --- | --- | --- |
|  |  |  | N (%) | N (% of DTC orders) | N | N (25%,75%) | N (% total tests logged) |
| **Chattanooga, TN** | April 1, 2021 | 41,200 | 14,423 (35.0) | 1,366 (9.5) | 2,722 | 1 (1, 3) | 1,900 (69.8) |
| **Ann Arbor/ Ypsilanti, MI** | June 4, 2021 | 20,000 | 10,115 (50.6) | 2,382 (23.5) | 8,198 | 1 (1, 3) | 6,344 (77.4) |
| **Fulton County, GA** | September 20, 2021 | 51,000 | 32,537 (63.8) | 2,012 (6.2) | 4,763 | 1 (1, 3) | 3,742 (78.6) |
| **O'ahu, HI** | September 19, 2021 | 125,000 | 79,536 (63.6) | 5,007 (6.3) | 17,140 | 2 (1, 4) | 11,154 (65.1) |
| **Indianapolis, IN** | October 18, 2021 | 35,300 | 22,970 (65.1) | 1,856 (8.1) | 5,724 | 2 (1, 4) | 5,199 (90.8) |
| **Louisville, KY** | October 11, 2021 | 40,500 | 19,204 (47.4) | 954 (5.0) | 2,918 | 2 (1, 4) | 2,626 (90.0) |
| **Total:** |  | 313,000 | 178,785 (57.1) | 14,398 (8.1) | 41,465 | 1 (1, 4) | 30,965 (75.0) |

<sup>a</sup>Percent of total kits distributed by direct-to-consumer (DTC) orders. Remainder of kits were distributed through in-person distribution.

### Reporting behaviors

Three-quarters (75.0%) of all tests logged in the digital assistant were reported to the state DoH (Figure 1, Supplemental Table 1). Sites with complete incentivization, Indiana and Kentucky, reported a higher proportion of test results to DoH than unincentivized or partially incentivized sites (p<0.001). The proportion of unreported results ranged from 9.2% (95% CI: 8.4-9.9) in Indiana to 34.9% (95% CI: 34.2-35.6) in Hawaii. In all intervention communities, positive results were less reported than negative results (60.4% vs 75.5%; p<0.001).

**Figure 1:**
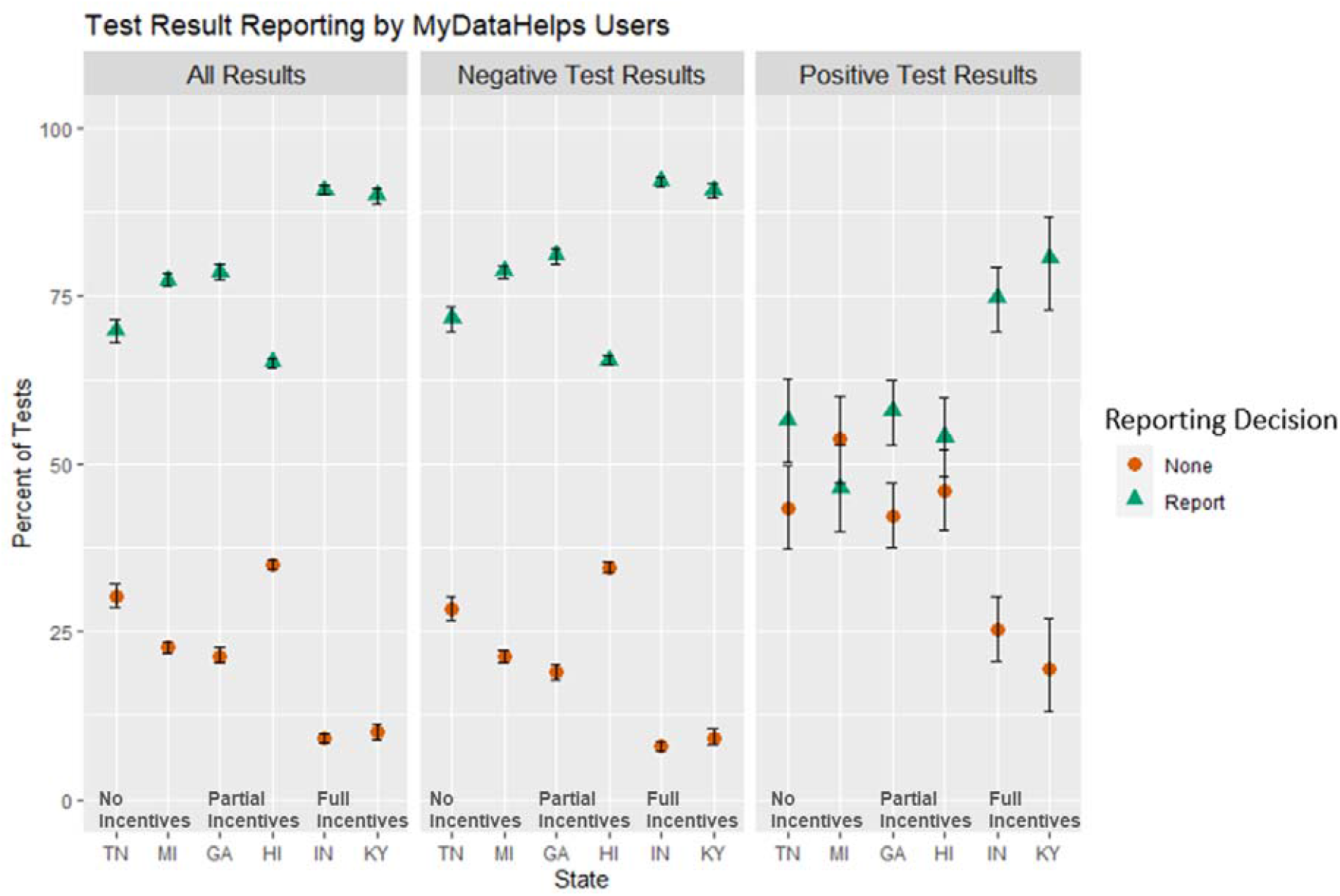
Reporting of Test Results among MyDataHelps Users by State.

## Discussion

Digital assistant users who logged test results made up a small proportion, only 8.1%, of households who ordered DTC test kits through the digital assistant. However, of those who logged test results, approximately 75% reported their results to their state’s DoH. The high usage of the digital assistant for ordering test kits demonstrates that the digital assistant was accessible to intervention communities. This implies that low usage of the digital assistant for logging tests may be due to inadequate community education about the importance of tracking and reporting home-test results. Further, there was a difference in reporting of tests based on result, with positive test results significantly less reported than negative results. It is important to understand and address the hesitations behind reporting positive tests.

The proportion of unreported tests was nearly three times higher in Tennessee and Hawaii compared to Indiana and Kentucky. This difference may be due to differences in incentivization structures, as participants in Indiana and Kentucky were incentivized to report tests throughout the intervention. Alternatively, test distribution in Indiana and Kentucky occurred in October 2021, following the Delta surge. Community awareness about the importance of reporting rapid antigen test results may have increased at this time relative to previous sites.

The high proportion of app users reporting their results to the DoH in Indiana and Kentucky suggests that app-based reporting systems may be successful in facilitating the reporting process when paired with incentives. However, the challenge remains drawing people to use the digital assistant, as evidenced by the low uptake of the digital assistant for testing purposes. Symptom-based participatory surveillance through digital applications has been used successfully for monitoring influenza-like-illness, among other infectious diseases, and rapid testing offers great opportunity to build on these technologies to rapidly ascertain changes in community prevalence of infection.^7,8^ Other means of improving uptake of the digital assistant or other reporting mechanisms should be explored further to maximize the value of these interventions.

### Strengths and Limitations

This report offers a unique look into COVID-19 test reporting behaviors of nearly fifteen thousand digital assistant users throughout the United States. However, there are limitations to this data. The number of digital assistant users is quite small compared to all intervention participants, and with the current data, we are unable to assess the demographics or socioeconomic status of digital assistant users, nor how digital assistant users compare to non-users.

### Conclusion

Three-quarters of those who used the digital assistant for testing reported their results to the DoH, indicating that app-based reporting may be an effective way to increase reporting of rapid tests for COVID-19. However, the relatively low voluntary uptake of the digital assistant indicates that user-centered strategies may be necessary to maximize digital assistant usage.

## Data Availability

All data produced in the present study are available upon reasonable request to the authors.

## Acknowledgments

We are deeply grateful first to the local communities of Louisville, KY; Indianapolis, IN; Fulton County, GA; O’ahu, HI; Ann Arbor/Ypsilanti, MI; and Chattanooga, TN, including the local health officials and second to our collaborators from the National Institute of Health (NIBIB and NHLBI) who provided scientific input into the design of this study and interpretation of our results, but could not formally join as co-authors due to institutional policies. We received meaningful contributions from Drs. Bruce Tromberg, Jill Heemskerk, Rachael Fleurence, Andrew Weitz, Krishna Juluru, Felicia Qashu, Dennis Buxton, Jue Chen and Erin Itturriaga.

---

**Supplemental Table 1:**

|  | **All Results** |  | **Negative Test Results** |  | **Positive Test Results** |  |
| --- | --- | --- | --- | --- | --- | --- |
|  | Reported<sup>a</sup><br>% (95% CI) | Not Reported<br>% (95% CI) | Reported<br>% (95% CI) | Not Reported<br>% (95% CI) | Reported<br>% (95% CI) | Not Reported<br>% (95% CI) |
| **Chattanooga, TN** | 69.8 (68-71.5) | 30.2 (28.5-32) | 71.7 (69.8-73.4) | 28.3 (26.6-30.2) | 56.5 (50.2-62.7) | 43.5 (37.3-49.8) |
| **Ann Arbor/ Ypsilanti, MI** | 77.4 (76.5-78.3) | 22.6 (21.7-23.5) | 78.7 (77.7-79.6) | 21.3 (20.4-22.3) | 46.3 (39.9-52.8) | 53.7 (47.2-60.1) |
| **Fulton County, GA** | 78.6 (77.4-79.7) | 21.4 (20.3-22.6) | 81.1 (79.9-82.2) | 18.9 (17.8-20.1) | 57.8 (52.9-62.5) | 42.2 (37.5-47.1) |
| **O'ahu, HI** | 65.1 (64.3-65.8) | 34.9 (34.2-35.6) | 65.5 (64.7-66.1) | 34.5 (33.8-35.3) | 54.0 (48.0-59.8) | 46.0 (40.2-52.0) |
| **Indianapolis, IN** | 90.8 (90.1-91.6) | 9.2 (8.4-9.9) | 92.1 (91.3-92.8) | 7.9 (7.2-8.7) | 74.8 (69.7-79.4) | 25.2 (20.6-30.3) |
| **Louisville, KY** | 90.0 (88.8-91.1) | 10.0 (8.9-11.2) | 90.8 (89.6-91.9) | 9.2 (8.1-10.4) | 80.6 (73.0-86.8) | 19.4 (13.2-27.0) |
| **Total** | 75.0 (74.6-75.4) | 25.0 (24.6-25.4) | 75.9 (75.4-76.3) | 24.1 (23.7-24.6) | 60.5 (58.1-62.8) | 39.6 (37.2-41.9) |

<sup>a</sup>Reported tests include those reported with full personal identifiable information and anonymously

